# Battle of Polio eradication in the Western Pacific Region in the transition to COVID-19 endemicity

**DOI:** 10.1101/2022.11.30.22282954

**Authors:** Ruobing Mei, Shirley L. L. Kwok, Eric H. Y. Lau, Tiffany H. K. Lo, Joseph T. Wu, Leesa K. Lin, Kathy Leung

## Abstract

The Polio eradication campaign has been set back substantially since 2020 due to the COVID-19 pandemic. Recent detections of poliovirus transmission in multiple high-income countries suggest suboptimal population immunity in many parts of the world even though polio vaccination has been included in routine childhood immunization for decades. We reviewed polio vaccination schedules and vaccine uptake in the Western Pacific Region countries and assessed the potential shortfall in population immunity against polio resurgence across these populations. In addition, we conducted a repeated cross-sectional study between 2021 and 2022 in the Western Pacific Region to understand factors contributing to polio vaccine hesitancy. Our results reveal potential shortfalls in population immunity against polio in Western Pacific Region and provide insights into how vaccination programs and campaigns can be strengthened to ensure continual progress towards polio eradication.

## Introduction

Despite great efforts being made in the progress of polio eradication over the last decade, the goal of achieving polio-free nations worldwide has taken longer than expected (1). Challenges remain in implementing the endgame strategies set by the Global Polio Eradication Initiative (GPEI), such as the shortages of resources for non-emergency health services resulting from the COVID-19 pandemic (2). As countries gradually loosen their COVID-19 travel restrictions, increased international travel poses risks of importing poliovirus. Furthermore, recent detections of circulating vaccine-derived polioviruses (cVDPVs) in London, New York State, and Israel reflect public health emergencies that call for prompt actions (3). The resurgence of polioviruses in high-income settings signals the potential for cVDPVs to spread globally, putting the unvaccinated population at high risk of contracting the paralytic disease.

Vaccines play a vital role in polio elimination as they effectively curb polioviruses and protect individuals from infection (4). Inactivated polio vaccine (IPV) and live attenuated oral polio vaccine (OPV) have been used worldwide to combat poliomyelitis. Although OPV provides strong intestinal immunity and halts transmission effectively, its weakened polioviruses could revert to active infectious form by reversing the attenuating nucleotide substitutions by positive selection pressure in the gastrointestinal tract (5–7), which causes vaccine-associated paralytic polio (VAPP) (8,9). Many developed countries have switched to IPV-only immunization schedules, following recommendations from the World Health Organization (WHO) (9). However, less developed countries are still in the process of introducing IPV in routine immunization programs due to manufacturing challenges and strong dependence on international aid (10). Under increased global mobility after the COVID-19 pandemic, cVDPVs would spread from OPV-using countries to countries that exclusively administer IPV, especially if the IPV vaccination coverage is not sufficiently high (11). To address the challenges of cVDPVs, high coverage of polio vaccination is desired to ensure population immunity to protect against polio.

Even though a wild polio-free status has been maintained in the Western Pacific Region for the past two decades, the threat of cVDPVs cannot be underestimated (12). In 2021, the polio vaccination coverages among 1-year-olds in Laos and the Philippines were 74% and 56%, which were below the 80% coverage target proposed by the World Health Organization (13). The considerable proportion of unvaccinated population and cross-border with countries with sporadic polio outbreaks call attention to potential cases of polio infection within the region. In this study, we reviewed the polio vaccination landscape in Western Pacific Region countries and evaluated potential gaps in population immunity. Furthermore, we conducted a study in nine Western Pacific Region countries to identify factors associated with polio vaccine hesitancy or refusal in order to aid the development of countermeasures.

### Polio Immunization Schemes and Vaccine Efficacy/Effectiveness

Following the global coordinated cessation of OPV to prevent the emergence of cVDPVs, high-income jurisdictions in Western Pacific Region, including Japan and South Korea, have adopted full IPV vaccination whereas most of their low- to middle-income counterparts such as Laos, Cambodia, Vietnam, and the Philippines are implementing various combination vaccination strategies (Figure 1) (14). Jurisdictions such as Hong Kong and Macau have introduced more than four doses of IPV in their routine childhood immunization program to consolidate the protection against polioviruses. Various parts of mainland China had begun to adopt sequential IPV and bivalent OPV (bOPV) schedules since December 2019, and some eastern coastal regions such as Shanghai and Jiangsu have switched to exclusive use of IPV (15,16).

**Figure 1.**
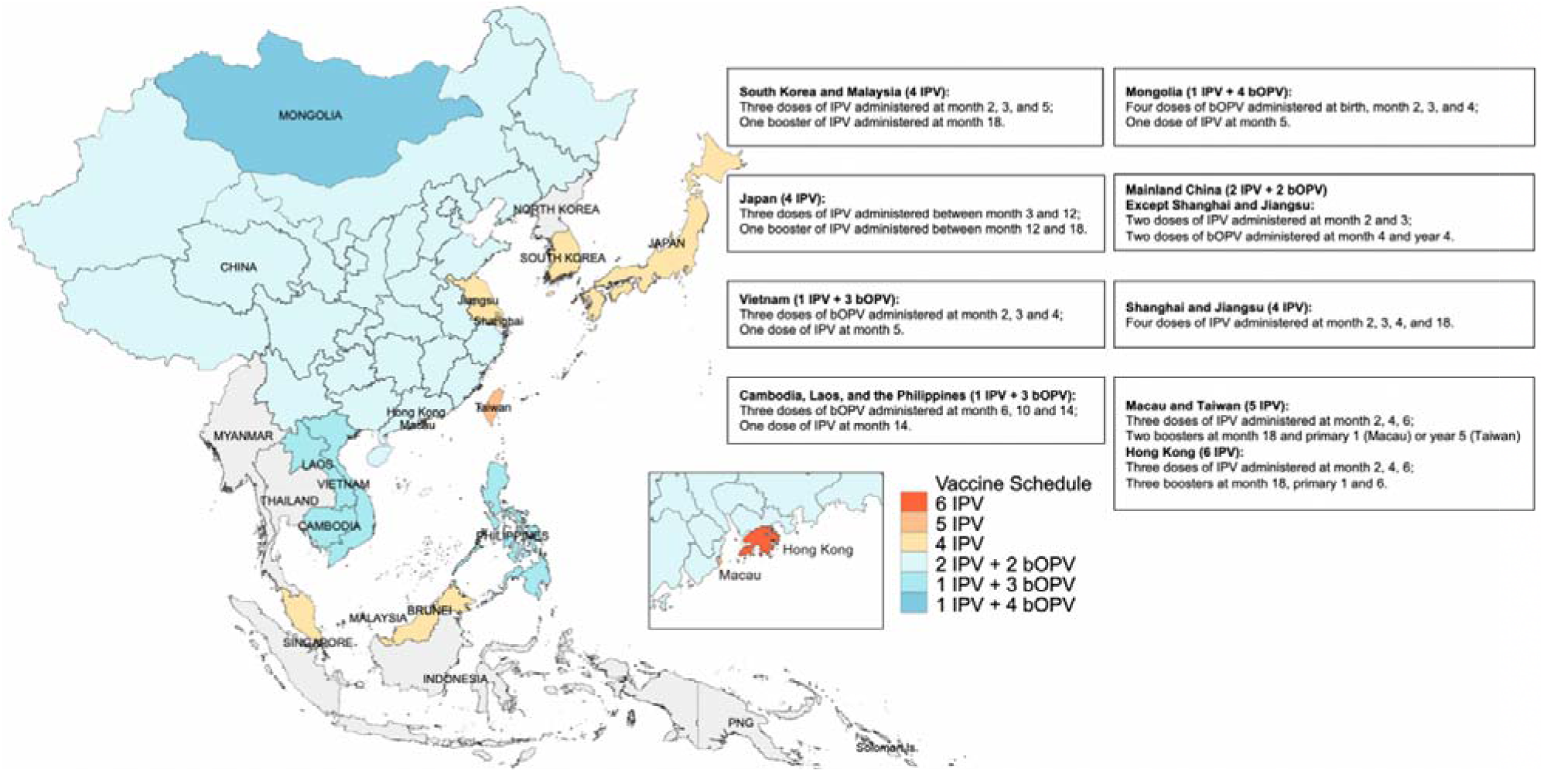
Poliovirus vaccination schedules in the Western Pacific Region countries or regions*. * Regions with lowercase letters denote provinces, municipality, or special administrative regions of China. Grey areas indicate countries or regions not included in our study. IPV = inactivated polio vaccine, bOPV = bivalent oral polio vaccine. Data Source: WHO vaccination schedules and Ministry of Health of Western Pacific Region countries (15,16,26–30).

We examined the efficacy and effectiveness of different polio vaccines by performing an evidence-based synthesis. A literature review of 26 studies of immunogenicity, efficacy and effectiveness of polio vaccines demonstrated that highly effective protection against polio infection was guaranteed with more than two doses of IPV or the co-administration of IPV and bOPV (Table S1). We considered seroconversion rate as a proxy measure of vaccine efficacy as it has been validated in previous studies (9,17): four doses of bOPV would reduce 94% of infection of serotypes 1 and 3 of polioviruses, whilst more than seven doses of bOPV would reduce 96% of polio infection (18–20). In contrast, more than two doses of IPV would reduce 96% of infection of all serotypes of polioviruses in the East Asian and American populations, and four doses of IPV series generally elicited 100% efficacy against poliomyelitis (21–23). There is no consensus with regard to the persistence of protection following polio vaccination, but scientific evidence shows that the protection could last for years (24,25).

### Critical Vaccination Coverage

In order to establish poliovirus population immunity and eradicate polio, it is crucial to understand the critical vaccination coverage required to get the basic reproduction number below 1. Using vaccine efficacy/effectiveness (VE) values from our literature review, we estimated a beta distribution for poliovirus VE; the details are provided in the supplementary text. We then estimated the critical vaccination coverage (V_c_) considering the basic reproduction number (R_0_) of poliovirus (31):

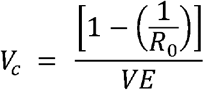

The critical coverage required ranges from 89·4% (95% CI: 80·0% to 100%) for a basic reproduction number of 5 to 95·7% (95% CI: 85·7% to 100%) for a basic reproduction number of 7. Compared to the countries’ actual vaccination coverages among 1-year-olds in 2021, Vietnam (81%), Laos (74%), and the Philippines’s (56%) vaccine uptakes fall under our estimated range of critical coverage required (Figure 2a and 2b).

**Figure 2a.**
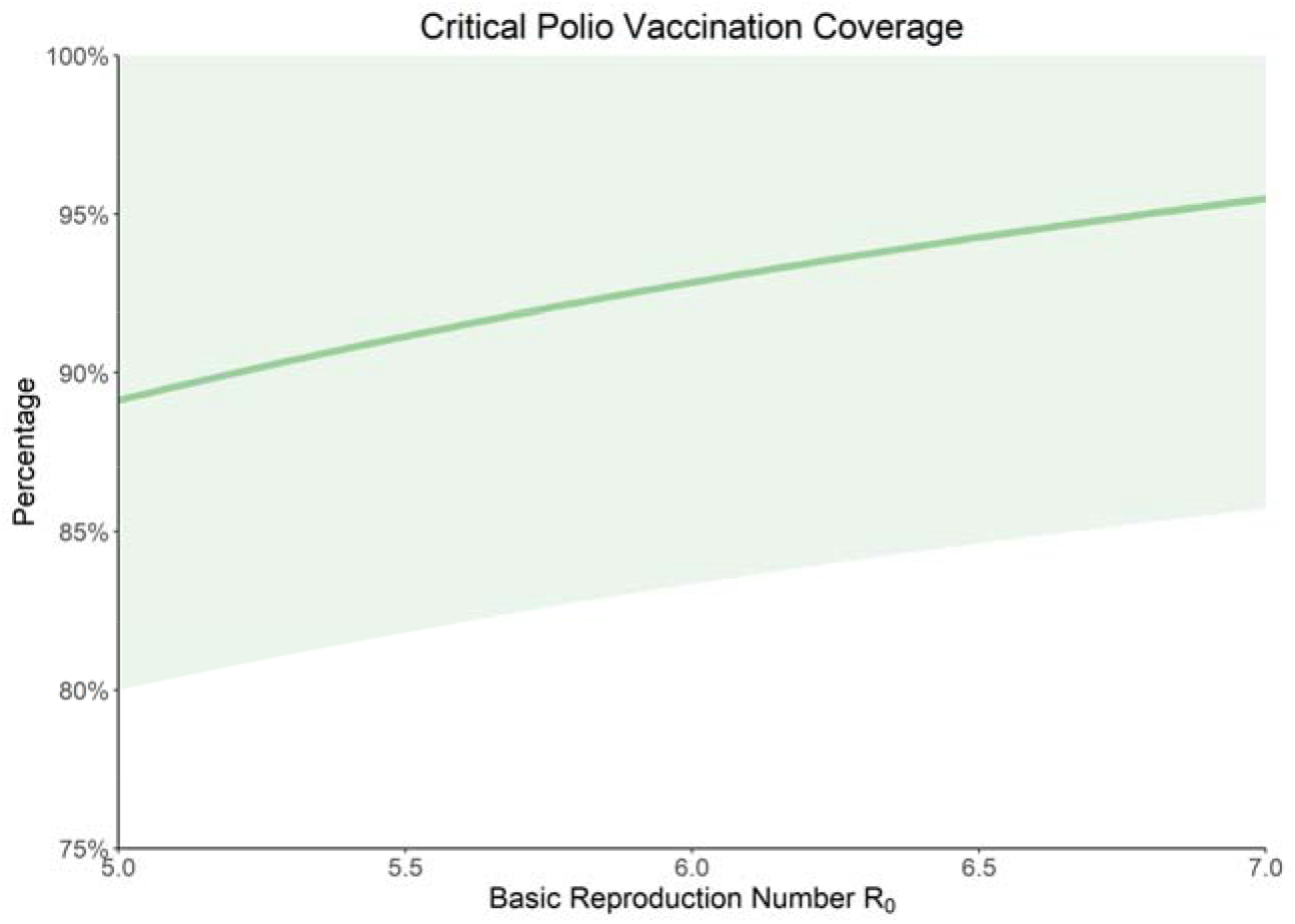
Estimation for the critical poliovirus vaccination coverage under varying poliovirus basic reproduction numbers*. * Shaded region represents 95% confidence interval for the critical vaccination coverage, estimates of vaccine effectiveness are detailed in Table S1.

**Figure 2b.**
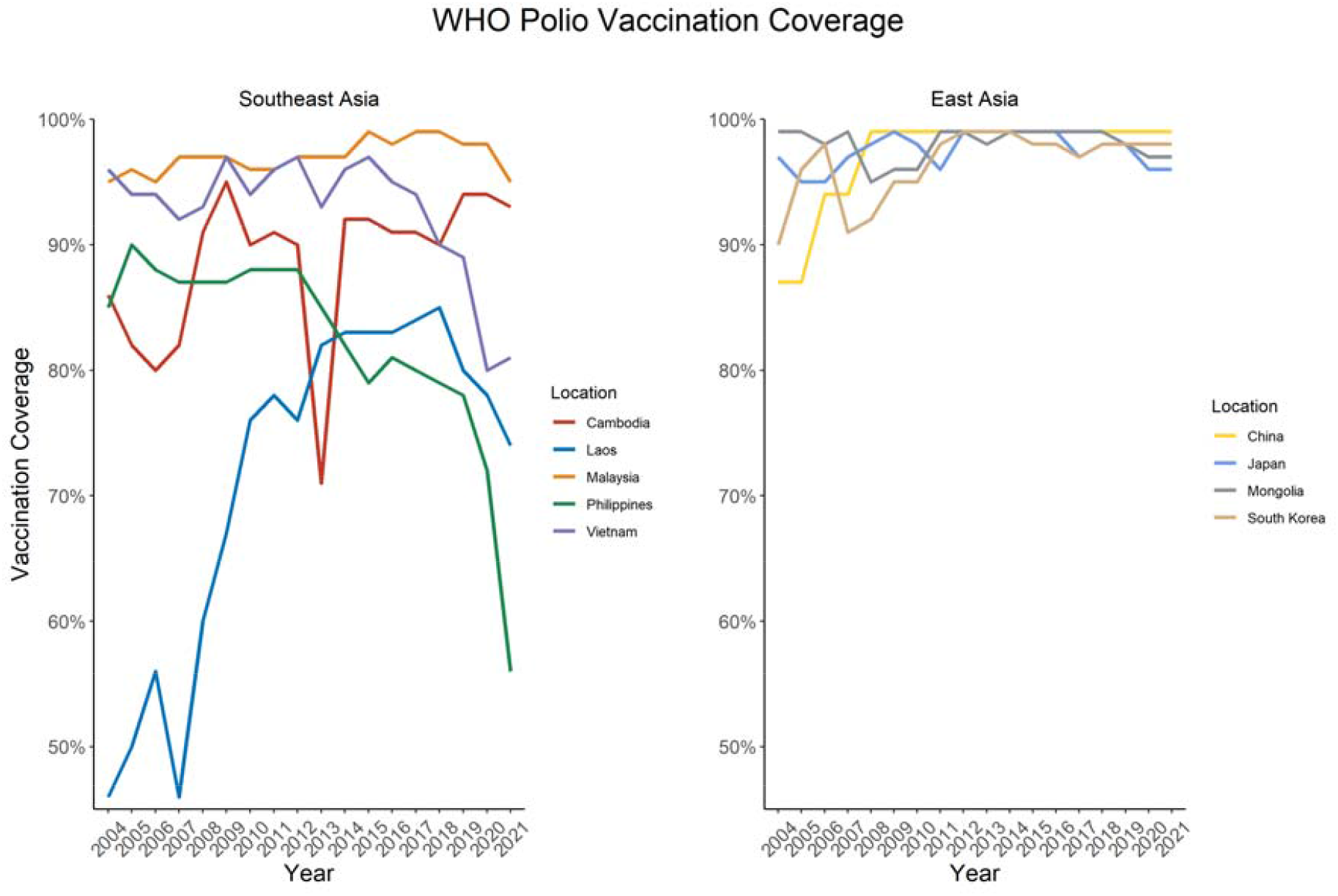
3^rd^ dose Polio vaccination (Pol3) coverage amongst 1-year-olds in Western Pacific Region countries, from 2004 to 2021*. * Data Source: 2004 to 2021 WHO polio (Pol3) vaccination coverage amongst 1-year-olds by country (32).

### Self-reported Vaccination Status

We conducted a repeated cross-sectional questionnaire-based survey in nine Western Pacific Region countries: Cambodia, China (mainland), Japan, Laos, Malaysia, Mongolia, the Philippines, South Korea, and Vietnam. Participants were recruited by ORB International, a monitoring and research agency (33), and provided consent prior to the interviews. Interviews were both web- and telephone-based and conducted in local languages. We conducted the first phase from June to August 2021, and the second phase from May to June 2022. The sample size for each phase was 2,000 for China and around 1,000 for other Asia-Pacific countries, with a margin of error below 1%. We interviewed 18,442 participants aged 18 years or older in the nine study locations. Our randomly selected sample is representative of the population in terms of gender, age, and region.

Participants were asked to report their knowledge, perceptions, and attitudes towards vaccination in addition to their sociodemographic information. 8,030 participants with children under the age of 18 also reported their child’s polio vaccination status. We considered the child to be vaccinated against polio if participants reported that their child had received at least one dose of polio vaccination or completed all government-recommended vaccinations. Compared to WHO’s official figures on polio vaccination coverage amongst 1-year-olds from 2004 to 2021, the self-reported uptake rates were lower in all nine countries, with Japan, Malaysia, Mongolia, and South Korea reporting over 50% below the official data (Figure 3). The self-reported vaccination coverages of children below 2 years old in Cambodia, China, Japan, Malaysia, and South Korea were lower than that of other age groups (Figure 3 and S1).

**Figure 3.**
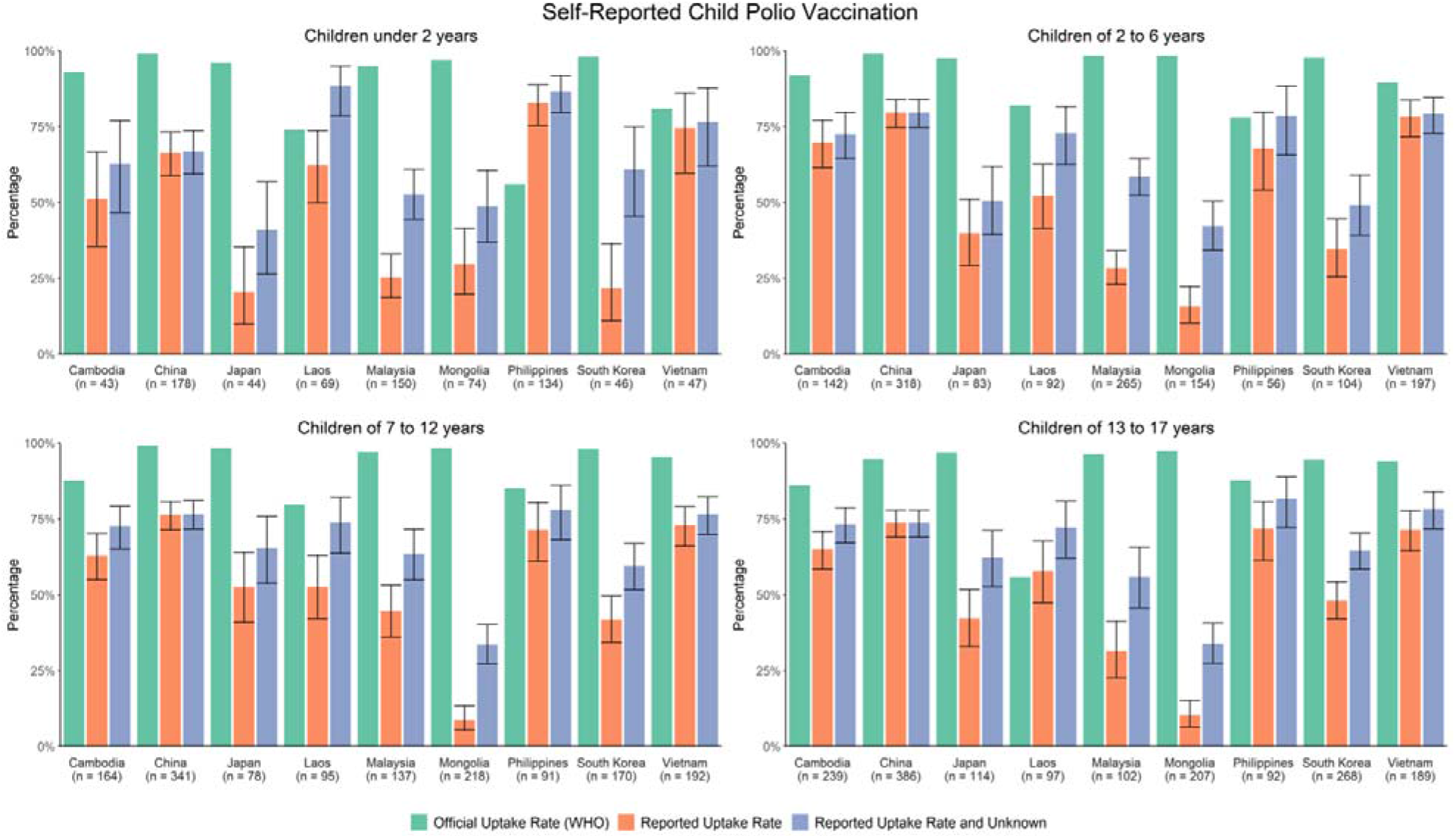
Official versus self-reported poliovirus vaccine uptake rate, by child age group*. * Self-reported uptake rate includes participants who either reported that their child had received any dose of polio vaccine or all recommended routine vaccinations; unknown includes participants who either reported that their child had received some vaccines but were not sure what they were or did not know if their child had been vaccinated or not. Children age is matched with WHO annual vaccination data. Data source: 2004 to 2021 WHO polio (Pol3) vaccination coverage amongst 1-year-olds by country (32).

### Factors Associated with Self-reported Polio Vaccination Status

We conducted binary logistic regression to examine factors associated with self-reported polio vaccination status. We used backward elimination process to select the covariates and finalize the model. The model controlled for country heterogeneity and time trend using child age as a proxy. Of those parents who reported that their children have not been vaccinated against polio, 59·0% reported that the COVID-19 pandemic made them more likely to get their children vaccinated against polio.

Several socioeconomic factors were associated with self-reported poliovirus vaccination status. The odds of children with self-reported vaccinations against polio were significantly influenced by and increased with their parents’ age, whereas no association was identified between self-reported polio vaccination status and children’s age. Compared with parents who had attained undergraduate education, those without formal education (adjusted OR = 0·64 [0·42-0·97], p = 0·037) were less likely to report that their children had been vaccinated against polio. Parental employment status was associated with self-reported childhood poliovirus vaccination status; the odds among parents who are students were 0.36 times that of employed parents (aOR = 0·36 [0·18-0·68], p = 0·002). Compared to Catholics, Buddhists (aOR = 0·64 [0·47-0·86], p = 0·003) and other religious groups (aOR = 0·62 [0·41-0·93], p = 0·022), including Hindu, Animism, Taoism, Jewish, and Confucianism were significantly less likely to report that their children had been vaccinated against polio. We included four Vaccine Confidence Index (VCI) statements in the model to examine whether parents’ perception of general vaccines determine self-reported child poliovirus vaccination status (34). Parents who perceived vaccines as being important for their children were significantly more likely to report that their children had been vaccinated against polio (aOR = 1·31 [1·04-1·66], p = 0·023). Accessibility of general vaccines (aOR = 1·21 [1·00-1·46], p = 0·046) was also associated with increased odds of self-reported vaccination against polio among parents.

**Table 1:**
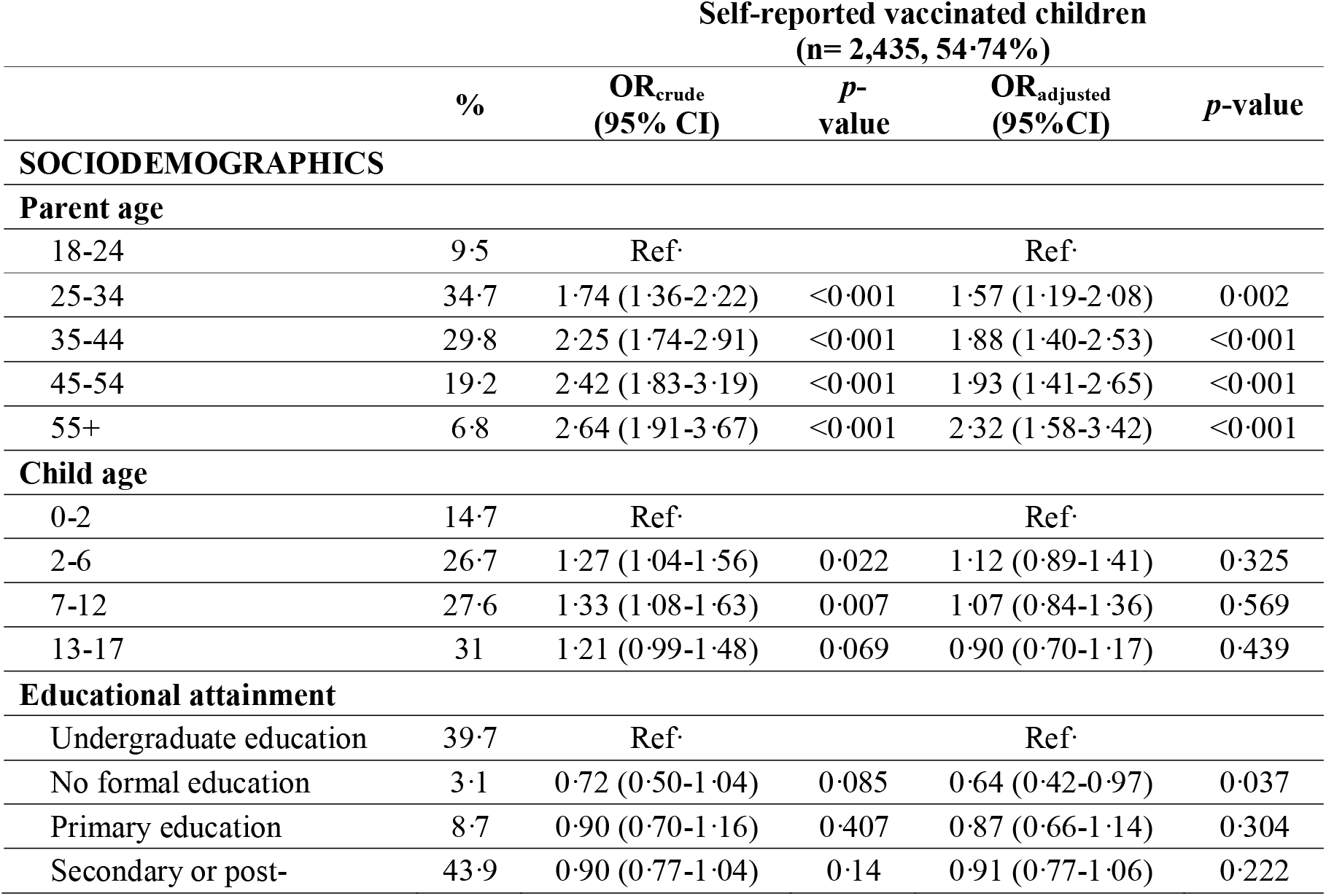
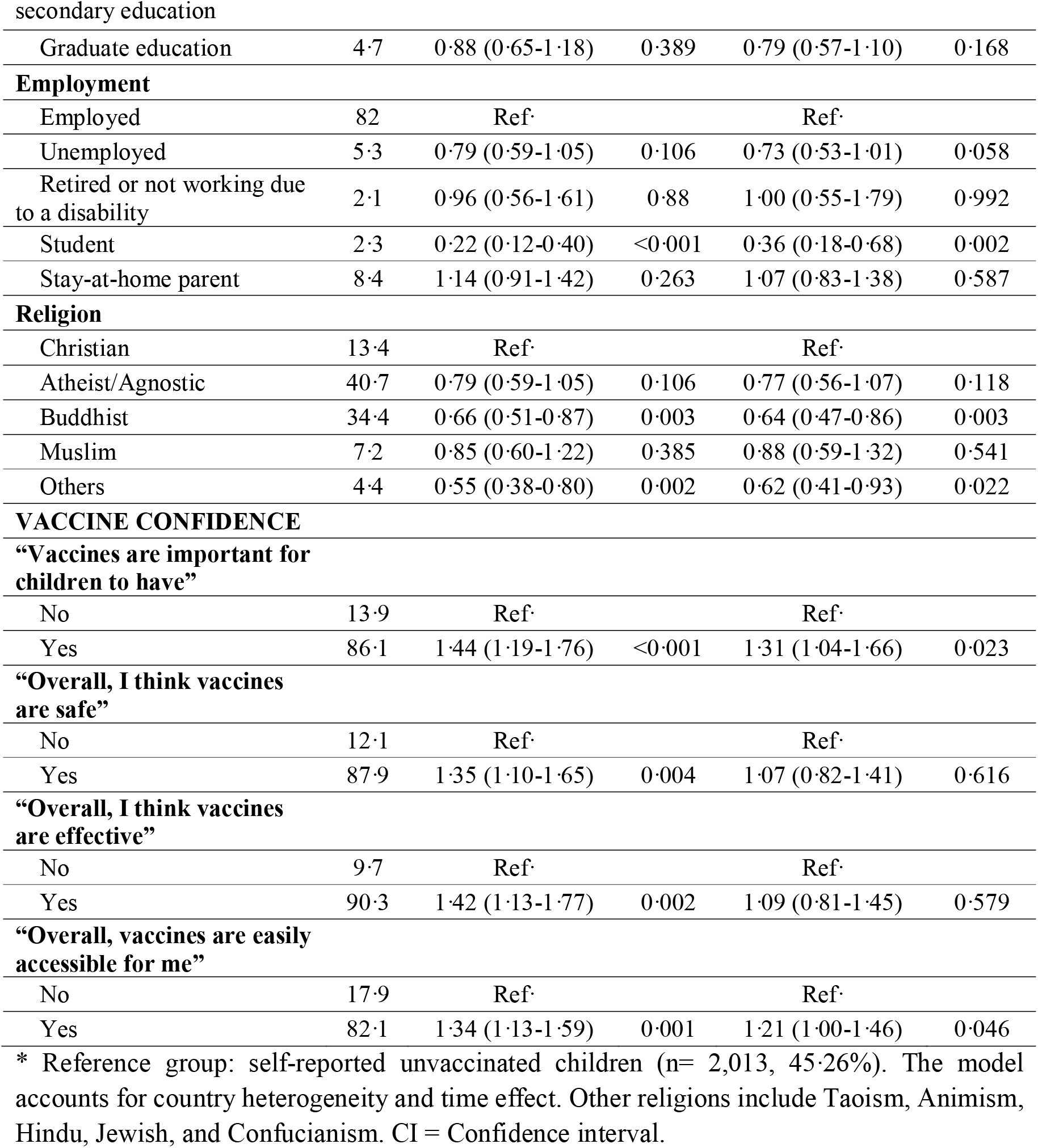
Factors associated with the self-reported uptake of polio vaccination in binary logistic regression model (n=4,448)*

## Discussion

With the resumption of international travel, the rise of vaccine hesitancy, and the resurgence of poliovirus cases around the globe, it is important to understand factors associated with reluctance to polio vaccination and close the immunization gap. Some Western Pacific Region countries are vulnerable due to both their high proportion of unvaccinated populations and interruptions in polio vaccination due to the COVID-19 pandemic. In particular, Laos, Vietnam, and the Philippines should put forth extra effort in order to achieve the critical vaccination coverage while other countries must maintain existing vaccination programs. The self-reported polio vaccine uptakes among Cambodian, Chinese, Japanese, Malaysian, and South Korean infants below two years old are lower than that of the older age groups, suggesting that parents of children under two years old are less aware of polio vaccination and there should be more focus on polio vaccine education amongst new parents.

The difference between self-reported poliovirus vaccine uptake and official figures could be explained by under-reporting. In particular, countries that use an online self-report questionnaire, namely Malaysia, South Korea, and Japan, all reported less than 50% of the official number; this is consistent with other online studies that also experienced under-reporting with online surveys (35–37). Another possible explanation is the lack of knowledge and health education of polio vaccines among the general public of these countries. This knowledge gap could increase the population’s vulnerability to vaccine misinformation and conspiracies, thus lowering vaccination intention and impeding governments’ immunization efforts as demonstrated in other studies (21,38–40).

We identify several sociodemographic factors associated with hesitancy in self-reported polio vaccine uptake, including parental age, educational level, employment status, and religious beliefs. Consistent with existing studies (41,42), older parents demonstrate less vaccine hesitancy for their children. Having an undergraduate degree is positively associated with self-reported polio vaccine acceptance compared to not having any formal education; this finding is in line with a previous study on routine childhood vaccination in the United States (43). The disparity in self-reported polio vaccination among parental educational levels could be explained by gaps in understanding vaccine risks and benefits. Besides, compared to those without formal education, parents who have completed undergraduate education tend to have a higher ability in information seeking and critical thinking after systematic educational training, which makes them less vulnerable to misinformation (44,45).

Our study finds that Buddhists and other small religious groups, including Hindu, Animist, Taoist, Jewish, and Confucianist, have a higher self-reported tendency towards polio vaccine refusal than Catholics. This result contradicts previous literature where Buddhists were generally found to have high vaccine acceptance (46–48), but is consistent with previous research conducted in Western Pacific Region (49–51). The vaccine hesitancy among Buddhists could be explained by the incompatibility between Buddhist belief to harm no living things and misinformation on polio vaccine production (48). In addition, Buddhists in Asia-Pacific tend to have high levels of religiosity due to the profound cultural and historical impacts of Buddhism in the region (52,53), while studies have shown that those individuals with strong religious faith often put less weight on authorities and health interventions (46,51,54). Since a sizable proportion of the Asia-Pacific population is Buddhist (55), it is important to establish efficient communication strategies between local health authorities and religious groups; the tailored communication should aim to improve understanding of polio vaccines, tackle misinformation, and promote vaccination. Besides, health authorities and non-governmental organizations should provide extensive training for community health workers and nurses on polio vaccine education, promotion, and vaccination outreach to increase public awareness and polio vaccine accessibility.

The results also reveal that parents’ perceptions of the importance of general vaccines for children influence self-reported child poliovirus vaccination status. Further communication about the importance of the polio vaccines for children should be addressed with parents, especially those with newborns. The accessibility of vaccines is also associated with self-reported polio vaccination of children by parents. As the production and supply of IPV remain a big challenge during the transition from OPV to IPV uses (41), local authorities should strengthen the collaboration with manufacturing and logistics companies to coordinate resource allocation and polio vaccine distribution. Our study further shows that more than half of the parents with self-reported non-vaccinated children have a higher willingness for polio vaccination after the COVID-19 pandemic. The re-emergence of poliovirus and elevated polio vaccination intention in the era of transitioning to COVID-19 endemicity reflect an appropriate time point to scale up polio vaccination in low-coverage areas to ensure population immunity against polio.

There are a few limitations to our study. First, the estimation of critical vaccination coverage in this study is based on a pool of polio vaccine efficacy and effectiveness for both IPV and bOPV over the globe due to the limited number of studies conducted in the Western Pacific Region, which might incur some inconsistency during the calculation. Second, our survey design might be prone to recall bias from survey respondents, hence resulting in low self-reported uptake rates. Third, the sample distributions of the countries based on online interviews are limited to the online population. The sampling method needs to be improved to ensure the representative of the general population. Nevertheless, our study serves as a tool for Western Pacific Region countries to adjust their target polio vaccination coverages and address the unvaccinated problem. Further research with more updated and country-specific measures is encouraged to provide more precise vaccination coverage estimates.

As polio eradication has been staggered during the COVID-19 pandemic and the gradual resumption of travel has made polioviruses a more imminent threat, governments and health authorities should address the unvaccinated issues as the willingness for polio vaccination during post-pandemic is running high. Other measures to tackle the polio vaccine misinformation should be incorporated simultaneously to increase vaccination intent and boost population immunity. Continuous surveillance, active engagement from stakeholders, and sufficient resources should be strategically put in place to prepare for the final battle of polio eradication after the COVID-19 pandemic.

## Data Availability

We collated data from publicly available secondary data sources. The sources of all secondary data included in the analyses are available in the main text or supplementary materials. Anonymised survey data will be made available to others upon request to the corresponding author with proposed data needs.

## Contributors

The study was conceived and curated by KL, LKL, and JTW, with important inputs from EHYL. LKL, THKL, RM, and SLLK collected data. RM and SLLK reviewed the literature, analysed data, and drafted the manuscript. KL led the study design and supervised the data analysis with EHYL. KL, LKL, JTW, EHYL, and THKL contributed to data interpretation, provided critical review, and commented on revisions of all text. All authors approved the final version of the manuscript.

## Ethics

This study was approved by the Institutional Review Board at the London School of Hygiene and Tropical Medicine (LSHTM 26636).

## Acknowledgements

The study is carried out with support from the Laboratory of Data Discovery for Health (D^2^4H). We gratefully acknowledge the help from Vaccine Confidence Project at the London School of Hygiene & Tropical Medicine and ORB International for providing relevant data. We also thank all survey participants for their important contributions to our study.

## References

1. Thompson KM, Duintjer Tebbens RJ. Lessons from the polio endgame: overcoming the failure to vaccinate and the role of subpopulations in maintaining transmission. The Journal of Infectious Diseases. 2017;216(suppl_1):S176–82.

2. Burkholder B, Wadood Z, Kassem AM, Ehrhardt D, Zomahoun D. The immediate impact of the COVID-19 pandemic on polio immunization and surveillance activities. Vaccine. 2021;

3. Polio eradication: falling at the final hurdle? The Lancet. 2022;400(10358):1079.

4. Lopalco PL. Wild and vaccine-derived poliovirus circulation, and implications for polio eradication. Epidemiology & Infection. 2017;145(3):413–9.

5. Cann AJ, Stanway G, Hughes PJ, Minor PD, Evans DM, Schild GC, et al. Reversion to neurovirulence of the live-attenuated Sabin type 3 oral poliovirus vaccine. Nucleic Acids Res. 1984 Oct 25;12(20):7787–92.

6. Famulare M, Chang S, Iber J, Zhao K, Adeniji JA, Bukbuk D, et al. Sabin Vaccine Reversion in the Field: a Comprehensive Analysis of Sabin-Like Poliovirus Isolates in Nigeria. J Virol. 2015 Oct 14;90(1):317–31.

7. Gnanashanmugam D, Falkovitz-Halpern MS, Dodge A, Fang M, Wong LJ, Esparza M, et al. Shedding and reversion of oral polio vaccine type 3 in Mexican vaccinees: comparison of mutant analysis by PCR and enzyme cleavage to a real-time PCR assay. J Clin Microbiol. 2007/06/20 ed. 2007 Aug;45(8):2419–25.

8. Wastewater monitoring comes of age. Nature Microbiology. 2022 Aug 1;7(8):1101–2.

9. Alfaro-Murillo JA, Ávila-Agüero ML, Fitzpatrick MC, Crystal CJ, Falleiros-Arlant LH, Galvani AP. The case for replacing live oral polio vaccine with inactivated vaccine in the Americas. The Lancet. 2020;395(10230):1163–6.

10. Bandyopadhyay AS, Orenstein WA. Evolution of inactivated poliovirus vaccine use for the endgame and beyond. Vol. 221, The Journal of Infectious Diseases. Oxford University Press US; 2020. p. 861–3.

11. Hill M, Bandyopadhyay AS, Pollard AJ. Emergence of vaccine-derived poliovirus in high-income settings in the absence of oral polio vaccine use. The Lancet. 2022;400(10354):713–5.

12. Kitamura K, Shimizu H. Outbreaks of Circulating Vaccine-Derived Poliovirus in the World Health Organization Western Pacific Region, 2000–2021. Japanese Journal of Infectious Diseases. 2022;75(5):431–44.

13. World Health Organization. Regional strategic framework for vaccine-preventable diseases and immunization in the Western Pacific 2021–2030. 2022;

14. Modlin JF, Bandyopadhyay AS, Sutter R. Immunization against poliomyelitis and the challenges to worldwide poliomyelitis eradication. The Journal of infectious diseases. 2021;224(Supplement_4):S398–404.

15. Jiangsu Commission of Health. Notification on adjustment of polio vaccination schedule in Jiangsu, China [Internet]. 2022 [cited 2022 Oct 3]. Available from: http://jspchfp.jiangsu.gov.cn/art/2022/9/23/art_7312_10613527.html

16. Shanghai Municipal Health Commission. Notification on adjustment of polio and measles vaccination schedule in Shanghai, China [Internet]. 2020 [cited 2022 Oct 3]. Available from: http://wsjkw.sh.gov.cn/gzdt1/20200728/d6440867ce9947898e463ba654c8a07f.html

17. Grassly NC. Immunogenicity and effectiveness of routine immunization with 1 or 2 doses of inactivated poliovirus vaccine: systematic review and meta-analysis. The Journal of infectious diseases. 2014;210(uppl_1):S439–46.

18. Sutter RW, Bahl S, Deshpande JM, Verma H, Ahmad M, Venugopal P, et al. Immunogenicity of a new routine vaccination schedule for global poliomyelitis prevention: an open-label, randomised controlled trial. The Lancet. 2015;386(10011):2413–21.

19. Saleem AF, Mach O, Yousafzai MT, Khan A, Weldon WC, Steven Oberste M, et al. Immunogenicity of different routine poliovirus vaccination schedules: a randomized, controlled trial in Karachi, Pakistan. The Journal of Infectious Diseases. 2018;217(3):443–50.

20. Chard AN, Martinez M, Matanock A, Kassem AM. Estimation of oral poliovirus vaccine effectiveness in Afghanistan, 2010–2020. Vaccine. 2021;39(42):6250–5.

21. Kim YK, Vidor E, Kim HM, Shin SM, Lee KY, Cha SH, et al. Immunogenicity and safety of a fully liquid DTaP-IPV-HB-PRP∼ T hexavalent vaccine compared with the standard of care in infants in the Republic of Korea. Vaccine. 2017;35(32):4022–8.

22. Nakayama T, Vidor E, Tsuzuki D, Nishina S, Sasaki T, Ishii Y, et al. Immunogenicity and safety of a DTaP-IPV/Hib pentavalent vaccine given as primary and booster vaccinations in healthy infants and toddlers in Japan. Journal of Infection and Chemotherapy. 2020;26(7):651–9.

23. Huoi C, Vargas-Zambrano J, Macina D, Vidor E. A combined DTaP-IPV vaccine (Tetraxim®/Tetravac®) used as school-entry booster: a review of more than 20 years of clinical and post-marketing experience. Expert Review of Vaccines. 2022;21(9):1215–31.

24. Larocca AMV, Bianchi FP, Bozzi A, Tafuri S, Stefanizzi P, Germinario CA. Long-Term Immunogenicity of Inactivated and Oral Polio Vaccines: An Italian Retrospective Cohort Study. Vaccines (Basel). 2022 Aug 17;10(8).

25. O’Grady M, Bruner PJ. Polio Vaccine. In: StatPearls [Internet]. StatPearls Publishing; 2021.

26. World Health Organisation. Vaccination schedule for Poliomyelitis by country [Internet]. Vaccination schedule for Poliomyelitis by country. [cited 2022 Oct 4]. Available from: https://immunizationdata.who.int/pages/schedule-by-disease/polio.html?ISO_3_CODE=&TARGETPOP_GENERAL=

27. Macau SAR Health Bureau. Macau SAR Immunization Programme -Recommended Children’s Schedule (from 2018) [Internet]. 2018 [cited 2022 Oct 3]. Available from: https://www.ssm.gov.mo/apps1/vaccine2/ch.aspx#clg11589

28. Hong Kong SAR Health Bureau. Hong Kong Childhood Immunisation Programme (from 2020) [Internet]. 2020 [cited 2022 Oct 3]. Available from: https://www.chp.gov.hk/files/pdf/updated_schedule_of_hkcip_recommended_by_scvpd_eng.pdf

29. Taiwan Centers for Disease Control. Taiwan Childhood Immunization Schedule [Internet]. 2019 [cited 2022 Oct 3]. Available from: https://www.cdc.gov.tw/Uploads/archives/3604cd18-8c64-4a49-a7c5-1b43df2f3f5b.pdf

30. Ministry of Health Malaysia. Malaysia Immunization Schedule for National Immunization Programme. [Internet]. [cited 2022 Oct 3]. Available from: http://www.myhealth.gov.my/en/immunisation-schedule/

31. Doherty M, Buchy P, Standaert B, Giaquinto C, Prado-Cohrs D. Vaccine impact: Benefits for human health. Vaccine. 2016 Dec 20;34(52):6707–14.

32. World Health Organisation. Poliomyelitis vaccination coverage: 2004-2021 [Internet]. Poliomyelitis vaccination coverage: 2004-2021. [cited 2022 Oct 11]. Available from: https://immunizationdata.who.int/pages/coverage/pol.html?CODE=WPR&YEAR=

33. ORB International [Internet]. ORB International - Global Research. [cited 2022 Nov 28]. Available from: https://orb-international.com/

34. Larson HJ, Schulz WS, Tucker JD, Smith DM. Measuring vaccine confidence: introducing a global vaccine confidence index. PLoS currents. 2015;7.

35. Dillman DA, Smyth JD, Christian LM. Internet, phone, mail and mixed-mode surveys: the tailored design method. Reis. 2016;154:161–76.

36. Andrade C. The Limitations of Online Surveys. Indian J Psychol Med. 2020 Nov;42(6):575–6.

37. Gao Z, House L, Xie J. Online Survey Data Quality and Its Implication for Willingness-to-Pay: A Cross-Country Comparison. Canadian Journal of Agricultural Economics/Revue canadienne d’agroeconomie. 2015 Apr 1;64.

38. Paul E, Steptoe A, Fancourt D. Attitudes towards vaccines and intention to vaccinate against COVID-19: Implications for public health communications. The Lancet Regional Health - Europe. 2021 Feb 1;1:100012.

39. Loomba S, de Figueiredo A, Piatek SJ, de Graaf K, Larson HJ. Measuring the impact of COVID-19 vaccine misinformation on vaccination intent in the UK and USA. Nature Human Behaviour. 2021 Mar;5(3):337–48.

40. Jolley D, Douglas KM. The effects of anti-vaccine conspiracy theories on vaccination intentions. PLoS One. 2014;9(2):e89177.

41. Lazarus JV, Ratzan SC, Palayew A, Gostin LO, Larson HJ, Rabin K, et al. A global survey of potential acceptance of a COVID-19 vaccine. Nature Medicine. 2021 Feb 1;27(2):225–8.

42. Temsah MH, Alhuzaimi AN, Aljamaan F, Bahkali F, Al-Eyadhy A, Alrabiaah A, et al. Parental attitudes and hesitancy about COVID-19 vs. routine childhood vaccinations: a national survey. Frontiers in public health. 2021;9:752323.

43. Kempe A, Saville AW, Albertin C, Zimet G, Breck A, Helmkamp L, et al. Parental Hesitancy About Routine Childhood and Influenza Vaccinations: A National Survey. Pediatrics. 2020 Jul;146(1).

44. Brandstetter S, Böhmer MM, Pawellek M, Seelbach-Göbel B, Melter M, Kabesch M, et al. Parents’ intention to get vaccinated and to have their child vaccinated against COVID-19: cross-sectional analyses using data from the KUNO-Kids health study. European journal of pediatrics. 2021;180(11):3405–10.

45. Wee MK, Cabantog J, Magpayo DD, Sabido NL, Samson E, David P. Factors causing vaccine hesitancy among parents in Bulacan. Studies in Medicine and Public Health. 2021;1(1):15–29.

46. Lahav E, Shahrabani S, Rosenboim M, Tsutsui Y. Is stronger religious faith associated with a greater willingness to take the COVID-19 vaccine? Evidence from Israel and Japan. The European Journal of Health Economics. 2022 Jun 1;23(4):687–703.

47. Thinane JS. Religious perspectives on Vaccination: Mandatory Covid-19 vaccine for SA Churches. Pharos Journal of Theology. 2022;103(10.46222).

48. Pelčić G, Karačić S, Mikirtichan GL, Kubar OI, Leavitt FJ, Cheng-Tek Tai M, et al. Religious exception for vaccination or religious excuses for avoiding vaccination. Croat Med J. 2016 Oct 31;57(5):516–21.

49. Larson HJ, de Figueiredo A, Xiahong Z, Schulz WS, Verger P, Johnston IG, et al. The State of Vaccine Confidence 2016: Global Insights Through a 67-Country Survey. EBioMedicine. 2016 Oct;12:295–301.

50. Du F, Chantler T, Francis MR, Sun FY, Zhang X, Han K, et al. Access to vaccination information and confidence/hesitancy towards childhood vaccination: a cross-sectional survey in China. Vaccines. 2021;9(3):201.

51. Brackstone K, Marzo RR, Bahari R, Head MG, Patalinghug ME, Su TT. COVID-19 Vaccine Hesitancy and Confidence in the Philippines and Malaysia: A Cross-sectional Study of Sociodemographic Factors and Information-Seeking. medRxiv. 2022;

52. Yeung GK, Chow W yin. ‘To take up your own responsibility’: the religiosity of Buddhist adolescents in Hong Kong. International Journal of Children’s Spirituality. 2010;15(1):5–23.

53. Saiyasak C. The Adaptation of Buddhism and Christianity to Asian Soils. In: ASM Strategic Mission Forum, Seoul, Korea. 2011.

54. Shelton RC, Snavely AC, De Jesus M, Othus MD, Allen JD. HPV vaccine decision-making and acceptance: does religion play a role? Journal of religion and health. 2013;52(4):1120–30.

55. Hackett C, Grim B, Stonawski M, Skirbekk V, Potančoková M, Abel G. The Global Religious Landscape: A Report on the Size and Distribution of the World’s Major Religious Groups as of 2010. 2012.

56. Sutter RW, Cochi SL. Inactivated Poliovirus Vaccine Supply Shortage: Is There Light at the End of the Tunnel? The Journal of Infectious Diseases. 2019 Oct 8;220(10):1545–6.

57. Wang Y, Xu Q, Jeyaseelan V, Ying Z, Mach O, Sutter R, et al. Immunogenicity of two-dose and three-dose vaccination schedules with Sabin inactivated poliovirus vaccine in China: an open-label, randomized, controlled trial. The Lancet Regional Health-Western Pacific. 2021;10:100133.

58. Yan S, Chen H, Zhang Z, Chang S, Xiao Y, Luo L, et al. Immunogenicity and safety of different sequential schedules of Sabin strain-based inactivated poliovirus vaccination: A randomized, controlled, open-label, phase IV clinical trial in China. Vaccine. 2020;38(40):6274–9.

59. Xiao S, Yang T, Fei Y, Deng P, Yang L. Basic immunogenicity and effectiveness of sequential immunization program of live attenuated and inactivated poliomyelitis vaccine. China Journal of Biologicals. 2020 Jul;33(7):809–12.

60. Kwak BO, Ma SH, Park SE, Shin SH, Choi KM, Lee TJ, et al. Comparison of the Immunogenicity and Safety of Three Enhanced Inactivated Poliovirus Vaccines from Different Manufacturers in Healthy Korean Infants: A Prospective Multicenter Study. Vaccines. 2020;8(2):200.

61. Chen H, Gao Z, Bai S, Liu X, Han S, Xiao Y, et al. Immunogenicity and safety of sabin-strain based inactivated poliovirus vaccine replacing salk-strain based inactivated poliovirus vaccine: An innovative application of different strain-IPVs replacement. Vaccine. 2021;39(17):2467–74.

62. Bandyopadhyay AS, Gast C, Rivera L, Sáez-Llorens X, Oberste MS, Weldon WC, et al. Safety and immunogenicity of inactivated poliovirus vaccine schedules for the post-eradication era: a randomised open-label, multicentre, phase 3, non-inferiority trial. The Lancet Infectious Diseases. 2021;21(4):559–68.

63. Ong-Lim AL, Shukarev G, Trinidad-Aseron M, Caparas-Yu D, Greijer A, Duchene M, et al. Safety and immunogenicity of 3 formulations of a Sabin inactivated poliovirus vaccine produced on the PER. C6® cell line: A phase 2, double-blind, randomized, controlled study in infants vaccinated at 6, 10 and 14 weeks of age. Human Vaccines & Immunotherapeutics. 2022;1–11.

64. Cuba IPV Study Collaborative Group. Randomized, placebo-controlled trial of inactivated poliovirus vaccine in Cuba. New England Journal of Medicine. 2007;356(15):1536–44.

65. O’Reilly KM, Durry E, ul Islam O, Quddus A, Abid N, Mir TP, et al. The effect of mass immunisation campaigns and new oral poliovirus vaccines on the incidence of poliomyelitis in Pakistan and Afghanistan, 2001–11: a retrospective analysis. The Lancet. 2012 Aug 4;380(9840):491–8.

66. Mangal TD, Aylward RB, Mwanza M, Gasasira A, Abanida E, Pate MA, et al. Key issues in the persistence of poliomyelitis in Nigeria: a case-control study. The lancet global health. 2014;2(2):e90–7.

67. Mahamud A, Kamadjeu R, Webeck J, Mbaeyi C, Baranyikwa MT, Birungi J, et al. Effectiveness of oral polio vaccination against paralytic poliomyelitis: a matched case-control study in Somalia. J Infect Dis. 2014 Nov 1;210 Suppl 1:S187–193.

68. Sutter RW, John TJ, Jain H, Agarkhedkar S, Ramanan PV, Verma H, et al. Immunogenicity of bivalent types 1 and 3 oral poliovirus vaccine: a randomised, double-blind, controlled trial. The Lancet. 2010;376(9753):1682–8.

69. Saleem AF, Mach O, Quadri F, Khan A, Bhatti Z, ur Rehman N, et al. Immunogenicity of poliovirus vaccines in chronically malnourished infants: a randomized controlled trial in Pakistan. Vaccine. 2015;33(24):2757–63.

70. Hu Y, Xu K, Han W, Chu K, Jiang D, Wang J, et al. Safety and immunogenicity of Sabin strain inactivated poliovirus vaccine compared with salk Strain Inactivated poliovirus vaccine, in different sequential schedules with bivalent oral poliovirus vaccine: randomized controlled noninferiority clinical trials in China. In: Open Forum Infectious Diseases. Oxford University Press US; 2019. p. ofz380.

71. He H, Wang Y, Deng X, Yue C, Tang X, Li Y, et al. Immunogenicity of three sequential schedules with Sabin inactivated poliovirus vaccine and bivalent oral poliovirus vaccine in Zhejiang, China: an open-label, randomised, controlled trial. The Lancet Infectious Diseases. 2020;20(9):1071–9.

72. Tagbo BN, Verma H, Mahmud ZM, Ernest K, Nnani RO, Chukwubike C, et al. Randomized Controlled Clinical Trial of Bivalent Oral Poliovirus Vaccine and Inactivated Poliovirus Vaccine in Nigerian Children. The Journal of infectious diseases. 2022;226(2):299–307.

