## Supplementary Materials for "Battle of Polio eradication in the Western Pacific Region in the transition to COVID-19 endemicity"

**Figure S1. Self-reported poliovirus vaccine uptake by child age group***

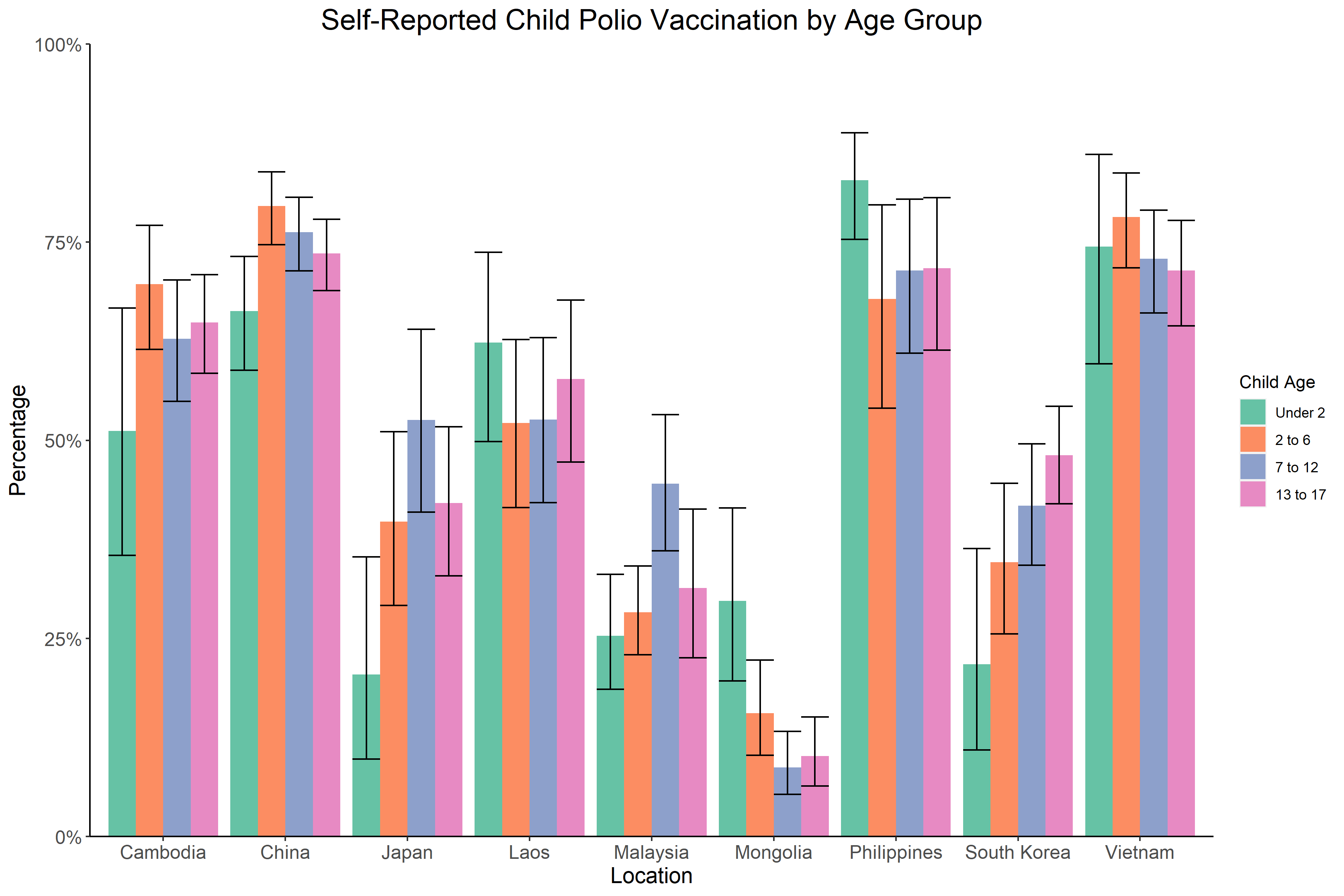

* Self-reported uptake rate includes participants who either reported that their child has received any dose of polio vaccine or all recommended routine vaccinations; unknown includes participants who either reported that their child has received some vaccines but were not sure what they were, or did not know if their child had been vaccinated or not.

**Figure S2. Estimated distribution of polio vaccine efficacy/effectiveness (VE)***

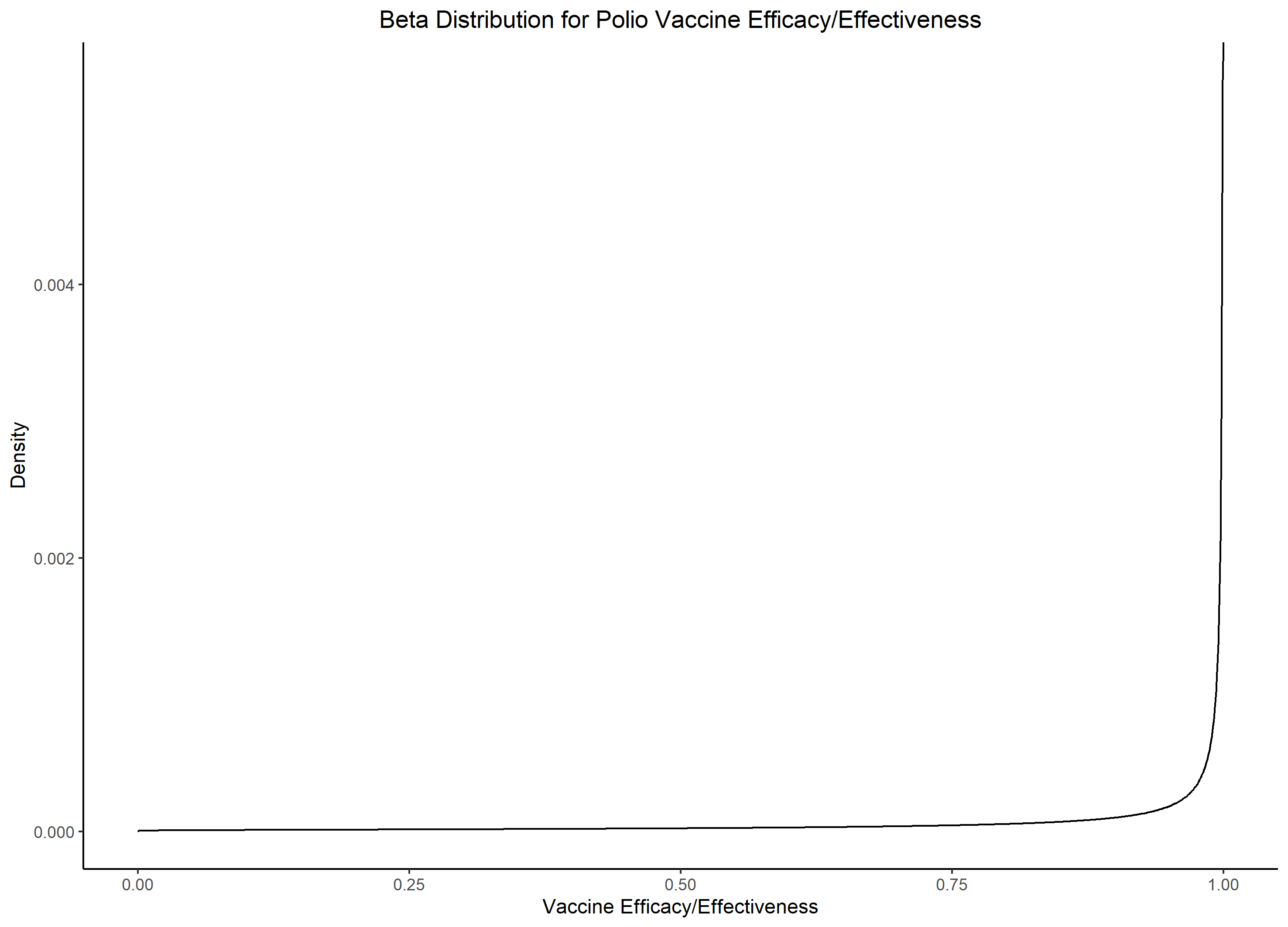

*Calculations are detailed in the supplementary text.

**Table S1. Vaccine efficacy/effectiveness (VE) of polio vaccines against paralytic poliomyelitis infection. IPV = inactivated polio vaccine, bOPV = bivalent oral polio vaccine, Hib = haemophilus influenza type b, HepB = hepatitis B, HB = hepatitis B.**

| ID | Vaccine Dosage & Type | Serotype of Polioviruses | VE | 95% CI | Study Location | Published Year | VE measure | Ref. |
| --- | --- | --- | --- | --- | --- | --- | --- | --- |
| Inactivated polio vaccine | |  |  |  |  |  |  |  |
| 1 | 2 IPV | Type 1 | 99·6% | 98·0-99·9% | China | 2021 | seroconversion | (57) |
|  |  | Type 2 | 97·1% | 94·3-98·5% |  |  |  |  |
|  |  | Type 3 | 100·0% | NA |  |  |  |  |
|  | 3 IPV | Type 1 | 100·0% | NA |  |  |  |  |
|  |  | Type 2 | 99·2% | 97·3-99·8% |  |  |  |  |
|  |  | Type 3 | 98·1% | 95·6-99·2% |  |  |  |  |
| 2 | 3 IPV | Type 1 | 100·0% | 98·1-100·0% | China | 2020 | seroconversion | (58) |
|  |  | Type 2 | 99·5% | 97·2-100·0% |  |  |  |  |
|  |  | Type 3 | 100·0% | 98·1-100·0% |  |  |  |  |
| 3 | 3 IPV | Type 1 | 100·0% | NA | China | 2020 | seroconversion | (59) |
|  |  | Type 3 | 98·6% | NA |  |  |  |  |
| 4 | 3 IPV (IPVAX) | Type 1 | 100·0% | NA | Korea | 2020 | seroconversion | (60) |
|  |  | Type 2 | 100·0% | NA |  |  |  |  |
|  |  | Type 3 | 100·0% | NA |  |  |  |  |
|  | 3 IPV (Imovax Polio) | Type 1 | 98·01% | NA |  |  |  |  |
|  |  | Type 2 | 96·2% | NA |  |  |  |  |
|  |  | Type 3 | 96·2% | NA |  |  |  |  |
|  | 3 IPV (Poliorix) | Type 1 | 98·3% | NA |  |  |  |  |
|  |  | Type 2 | 100·0% | NA |  |  |  |  |
|  |  | Type 3 | 100·0% | NA |  |  |  |  |
| 5 | 3 IPV | Type 1 | 100·0% | 97·8-100·0% | China | 2021 | seroconversion | (61) |
|  |  | Type 2 | 100·0% | 97·8-100·0% |  |  |  |  |
|  |  | Type 3 | 100·0% | 97·8-100·0% |  |  |  |  |
| ID | **Vaccine Dosage & Type** | **Serotype of Polioviruses** | **VE** | **95% CI** | **Study Location** | **Published Year** | **VE indicator** | **Ref.** |
| 6 | 3 IPV | Type 1 | 100·0% | 97·9-100·0% | Panama & the Dominican Republic | 2021 | seroconversion | (62) |
|  |  | Type 2 | 100·0% | 97·9-100·0% |  |  |  |  |
|  |  | Type 3 | 100·0% | 97·9-100·0% |  |  |  |  |
| 7 | 3 IPV | All | 99·0-100·0% | NA | NA |  | clinical efficacy/ effectiveness | (25) |
| 8 | 3 IPV | Type 1 | 89·0% | 79·5-95·5% | Philippines | 2022 | seroconversion | (63) |
|  |  | Type 2 | 98·0% | 92·5-99·0% |  |  |  |  |
|  |  | Type 3 | 100·0% | 95·0-100·0% |  |  |  |  |
| 9 | 4 IPV | Type 1 | 80·0% | 72·0-86·0% | Pakistan | 2018 | seroconversion | (19) |
|  |  | Type 2 | 84·0% | 78·0-90·0% |  |  |  |  |
|  |  | Type 3 | 93·0% | 89·0-98·0% |  |  |  |  |
| 10 | 4 DPT-IPV | Type 1 | 100·0% | 98·2-100·0% | Japan | 2020 | seroconversion | (22) |
|  |  | Type 2 | 100·0% | 98·2-100·0% |  |  |  |  |
|  |  | Type 3 | 100·0% | 98·2-100·0% |  |  |  |  |
| 11 | 3 DPT-Hib-IPV | Type 1 | 94·0% | 84·0-99·0% | Cuba | 2007 | seroconversion | (64) |
|  |  | Type 2 | 83·0% | 70·0-92·0% |  |  |  |  |
|  |  | Type 3 | 100·0% | 94·0-100·0% |  |  |  |  |
| 12 | 3 DTaP-IPV-HepB-Hib (Pentaxim) | Type 1 | 100·0% | 97·2-100·0% | South Korea | 2017 | seroconversion | (21) |
|  |  | Type 2 | 100·0% | 97·2-100·0% |  |  |  |  |
|  |  | Type 3 | 100·0% | 97·2-100·0% |  |  |  |  |
|  | 4 DTaP-IPV//PRP∼T+HB (Hexaxim) | Type 1 | 100·0% | 97·2-100·0% |  |  |  |  |
|  |  | Type 2 | 100·0% | 97·2-100·0% |  |  |  |  |
|  |  | Type 3 | 100·0% | 97·2-100·0% |  |  |  |  |
| ID | **Vaccine Dosage & Type** | **Serotype of Polioviruses** | **VE** | **95% CI** | **Study Location** | **Published Year** | **VE indicator** | **Ref.** |
| Oral polio vaccine | |  |  |  |  |  |  |  |
| 13 | 1 bOPV | All | 23·4% | 10·4-34·6% | Pakistan and Afghanistan | 2012 | clinical effectiveness | (65) |
| 14 | 1 bOPV | Type 1 | 29·5% | 20·1-38·4% | Nigeria | 2014 | clinical efficacy | (66) |
|  |  | Type 3 | 23·8% | 5·3-44·9% |  |  |  |  |
| 15 | 1-3 bOPV | All | 92·0% | 72·0-98·0% | Somalia | 2014 | clinical effectiveness | (67) |
|  | 4 bOPV | All | 97·0% | 89·0-99·0% |  |  |  |  |
| 16 | 2 bOPV | Type 1 | 86·0% | 79·0-90·0% | India | 2010 | seroconversion | (68) |
|  |  | Type 2 | 11·0% | 7·0-17·0% |  |  |  |  |
|  |  | Type 3 | 74·0% | 66·0-80·0% |  |  |  |  |
| 17 | 4 bOPV | Type 1 | 98·7% | 95·4-99·8% | India | 2015 | seroconversion | (18) |
|  |  | Type 2 | 18·7% | 12·9-25·7% |  |  |  |  |
|  |  | Type 3 | 97·4% | 93·5-99·3% |  |  |  |  |
| 18 | 4 bOPV | Type 1 | 97·0% | 93·0-100·0% | Pakistan | 2018 | seroconversion | (19) |
|  |  | Type 2 | 19·0% | 13·0-27·0% |  |  |  |  |
|  |  | Type 3 | 94·0% | 90·0-98·0% |  |  |  |  |
| 19 | 1 bOPV |  | 32·0% | 24·0-39·0% | Afghanistan | 2021 | clinical efficacy | (20) |
|  | ≥7 bOPV |  | 96·0% | 90·0-99·0% |  |  |  |  |
| Combination schedule | |  |  |  |  |  |  |  |
| 20 | 1 IPV + 1 bOPV | Type 1 | 93·8% | 69·8-99·8% | Pakistan | 2015 | seroconversion | (69) |
|  |  | Type 2 | 100·0% | NA |  |  |  |  |
|  |  | Type 3 | 93·5% | NA |  |  |  |  |
| 21 | 1 IPV + 1 bOPV | Type 1 | 99·4% | 96·5-100·0% | India | 2015 | seroconversion | (18) |
|  |  | Type 2 | 68·6% | 60·7-75·8% |  |  |  |  |
|  |  | Type 3 | 99·4% | 96·5-100·0% |  |  |  |  |
|  | 2 IPV + 1 bOPV | Type 1 | 99·4% | 96·5-100·0% |  |  |  |  |
|  |  | Type 2 | 78·1% | 70·7-84·3% |  |  |  |  |
|  |  | Type 3 | 98·7% | 95·4-99·8% |  |  |  |  |
| ID | **Vaccine Dosage & Type** | **Serotype of Polioviruses** | **VE** | **95% CI** | **Study Location** | **Published Year** | **VE indicator** | **Ref.** |
| 22 | 1 IPV + 2 bOPV | Type 1 | 100·0% | 96·6-100·0% | China | 2019 | seroconversion | (70) |
|  |  | Type 2 | 83·2% | 74·7-89·7% |  |  |  |  |
|  |  | Type 3 | 100·0% | 96·6-100·0% |  |  |  |  |
|  | 2 IPV + 1 bOPV | Type 1 | 99·2% | 95·4-100·0% |  |  |  |  |
|  |  | Type 2 | 94·9% | 93·6-99·8% |  |  |  |  |
|  |  | Type 3 | 100·0% | 96·7-100·0% |  |  |  |  |
| 23 | 1 IPV + 2 bOPV | Type 1 | 100·0% | NA | China | 2020 | seroconversion | (71) |
|  |  | Type 2 | 62·0% | NA |  |  |  |  |
|  |  | Type 3 | 100·0% | NA |  |  |  |  |
|  | 2 IPV + 1 bOPV | Type 1 | 100·0% | NA |  |  |  |  |
|  |  | Type 2 | 95·0% | NA |  |  |  |  |
|  |  | Type 3 | 100·0% | NA |  |  |  |  |
| 24 | 1 IPV + 2 bOPV | Type 1 | 100·0% | 98·1-100·0% | China | 2020 | seroconversion | (58) |
|  |  | Type 2 | 91·5% | 86·6-95·1% |  |  |  |  |
|  |  | Type 3 | 100·0% | 98·1-100·0% |  |  |  |  |
|  | 2 IPV + 1 bOPV | Type 1 | 100·0% | 98·0-100·0% |  |  |  |  |
|  |  | Type 2 | 98·4% | 95·3-99·67% |  |  |  |  |
|  |  | Type 3 | 100·0% | 98·0-100·0% |  |  |  |  |
| 25 | 1IPV + 4 bOPV | Type 1 | 94·0% | 90·0-98·0% | Pakistan | 2017 | seroconversion | (19) |
|  |  | Type 2 | 53·0% | 44·0-61·0% |  |  |  |  |
|  |  | Type 3 | 98·0% | 95·0-100·0% |  |  |  |  |
| 26 | 2 IPV + 4 bOPV | Type 1 | 98·9% | 96·7-99·8% | Nigeria | 2020 | seroconversion | (72) |
|  |  | Type 2 | 95·9% | 92·8-97·9% |  |  |  |  |
|  |  | Type 3 | 98·1% | 88·2-94·8% |  |  |  |  |
|  | 3 IPV + 1 bOPV | Type 1 | 89·6% | 85·4-93·0% |  |  |  |  |
|  |  | Type 2 | 93·7% | 90·1-96·3% |  |  |  |  |
|  |  | Type 3 | 98·5% | 96·3-99·6% |  |  |  |  |

**Supplementary Text: Estimation of poliovirus efficacy/effectiveness**

Using vaccine efficacy/effectiveness (VE) values from our literature review, we estimated a beta distribution for poliovirus VE (both IPV and bOPV), where the distribution is:

$$f\left( x | \alpha,\beta\right) = \frac{x^{\alpha-1}\left( 1-x \right)^{\beta-1}}{\frac{\Gamma\left( \alpha\right)\Gamma\left( \beta\right)}{\Gamma\left( \alpha+\beta\right)}}$$

where $\Gamma$ is the Gamma function, and $\alpha$ and $\beta$ are as follow:

$\alpha= \left( \frac{1-\mu}{\sigma^{2}}-\frac{1}{\mu} \right)\mu^{2}$, and

$$\beta= \alpha\left( \frac{1}{\mu}-1 \right)$$
